# Correlates of COVID-19 vaccine acceptance, hesitancy and refusal among employees of a safety net California county health system with an early and aggressive vaccination program: Results from a cross-sectional survey

**DOI:** 10.1101/2021.09.14.21263588

**Authors:** Nicole M. Gatto, Jerusha E. Lee, Donatella Massai, Susanna Zamarripa, Bijan Sasaninia, Dhruv Khurana, Kelsey Michaels, Deborah Freund, Judi Nightingale, Anthony Firek, for the Riverside University Health System (RUHS) Comparative Effectiveness and Clinical Outcomes Research Center (CECORC)-Claremont Graduate University (CGU) COVID-19 Research Group

## Abstract

Information on vaccine acceptance among healthcare workers is needed as health professionals provide front line care to COVID-19 patients. We developed and implemented an anonymous internet-based cross-sectional survey with direct solicitation among employees of a safety net health system. Items queried demographic and health-related characteristics, experience with and knowledge of COVID-19, and determinants of decisions to vaccinate. COVID-19 vaccine acceptance groups (acceptors, hesitant, refusers) were defined; an adapted version of the WHO vaccine hesitancy scale was included. The survey demonstrated good reliability (Cronbach’s alpha = 0.92 for vaccine hesitancy scale; 0.93 for determinants). General linear and logistic regression methods examined factors which were univariately associated with vaccine hesitancy and vaccine acceptance, respectively. Multivariable models were constructed with stepwise model-building procedures. Race/ethnicity, marital status, job classification, immunocompromised status, flu vaccination and childhood vaccination opinions independently predicted hesitancy scale scores. Gender, education, job classification and BMI independently predicted acceptance, hesitancy and refusal groups. Among hesitant employees, uncertainty was reflected in reports of motivating factors influencing their indecision. Despite a strong employee-support environment and job protection, respondents reported physical and mental health effects. Appreciation of varied reasons for refusing vaccination should lead to culturally sensitive interventions to increase vaccination rates in healthcare workers.

## 1. Introduction

The success of any vaccination program is dependent on a number of interconnected and interdependent actions. These include development of vaccine, testing for efficacy and safety, rapid distribution to the population and acceptance by recipients. The latter issue of vaccine uptake [1] is critical, and can be characterized as vaccine acceptance, refusal or hesitancy. Vaccine hesitancy is formally defined as a “delay in acceptance or refusal of vaccines despite availability of vaccine services”[2], and is acknowledged as complex and context-specific with the potential to vary across time and place. Factors that may influence vaccine acceptance include complacency, access to vaccination sources, confidence in the safety and efficacy and perceived intent of the agencies providing the vaccine[3].

Historically, themes have characterized perspectives among individuals who are vaccine hesitant in the United States (US)[4-6] and are not mutually exclusive. One is a distrust of government and a desire to retain individual liberties including the right to make their own decisions versus governmental-imposed actions [4,6]. Second, are philosophical beliefs that stress the purity of the human body [7,8], and further, are fears and controversies related to vaccine safety [6]. The spread of the more infectious SARS-CoV-2 delta variant in the US and the surge in new cases mainly in the unvaccinated demonstrates the liability of personal decisions on vaccine acceptance. Vaccine hesitancy is global and not limited to the current COVID-19 pandemic; it has, in part, been blamed for international resurgences of infectious disease outbreaks such as measles, which had previously been brought under control [7].

Since the emergence of the SARS-CoV-2 virus in late 2019 in Wuhan, China, there have been an excess of 216 million confirmed cases and 4.5 million deaths globally[9], with the US having the highest number of deaths at approximately 640,000 as of September 1, 2021 [10]. The pandemic has strained public health and medical systems, caused severe illness and death among a proportion of those infected overwhelming hospitals and healthcare workers, and caused major disruptions of daily life[11]. While non-pharmaceutical public health interventions (NPIs) aimed at slowing the spread of SARS-CoV-2 were key components of the early pandemic response, the availability of vaccines against COVID-19 beginning in January 2021 in the US has enhanced and strengthened primary prevention strategies. As such, vaccine uptake among populations eligible for vaccination is essential to ending the pandemic.

A consequence of the rapid development of successful vaccines during the COVID-19 pandemic has brought into refocus the issue of vaccine hesitancy, which the WHO has previously identified as one of the top ten global health threats of 2019 [12]. Surveys of the general US population between November 2020 and March 2021 prior to when the current study was conducted showed that the proportion who plan to be vaccinated has increased during this time. However, depending on the survey, 14-17% have expressed hesitancy and 10-15% report that they definitely won’t get vaccinated[13-16]. This raises concerns as to whether reaching herd immunity in the US will be achievable.

While the US population has been surveyed recurrently and assessments provide insights into factors influencing a person’s decision to accept vaccination [17-21], fewer studies of COVID-19 vaccine hesitancy have focused on healthcare workers and health system employees [22-24]. One survey between September and October 2020 found that 35% of health system employees expressed apprehension concerning the possibility of serious adverse events from vaccines, with 67% reporting they would delay COVID-19 vaccination if a vaccine became available[22]. A Kaiser Family Foundation/Washington Post survey in March 2021 indicated that 48% of healthcare workers surveyed had not been vaccinated with 12% undecided about vaccination and 18% not planning to get vaccinated[25]. A review of studies among healthcare workers which included several from the US identified common reasons provided by hesitant employees [26]. However, all surveys were conducted prior to vaccines becoming available in the US or within the first month after they were administered and suggested that the fraction of the population that is hesitant may have decreased over the one-year period covered by the review, indicating as did another study [27] that timing may have had an effect on hesitancy.

Information on vaccine acceptance, hesitancy and refusal among healthcare workers is needed as doctors, nurses, physician assistants, pharmacists and other health professionals provide front line care to COVID-19 patients. Frontline workers directly interact with patients, thereby having a high potential for exposure to the SARS-CoV-2 virus, emphasizing the essential need for protection through vaccination. In fact, healthcare workers were among the first groups to be eligible for COVID-19 vaccination in the US and California [11]. Additionally, health professionals serve as a direct and trusted source of information for patients [28], raising the question as to whether their opinions on vaccination could indirectly influence the medical advice they provide. Illustrating this concern, research among French physicians demonstrated high vaccination hesitancy translated into lower vaccine recommendations to their patients [29].

There is a present and unmet need to better understand vaccine uptake in healthcare workers and influences underlying their decisions. This is critical so that actions can be taken to both persuade them to be vaccinated and to retain their employment given potential shortages during times of spikes in the demand for hospital care. In response to this need, many states have begun to mandate vaccination among health workers [30]. As determined by earlier studies, vaccine acceptance or refusal among healthcare workers may or may not have distinct determinants from those for other populations and other types of vaccines [23,26,31]. Understanding determinants among healthcare workers and health system employees could lead to more focused worker- and patient-centered educational and other interventions, as employment retention and achieving herd immunity are critical to “ending” COVID-19 pandemic in these populations. This is particularly true in safety net medical centers that treat the most vulnerable who are at higher risk for COVID-19 complications and death. The Riverside University Health System (RUHS) medical center serves a large and highly disadvantaged predominantly multi-ethnic population. An early and aggressive COVID-19 vaccination program was initiated that virtually assured any employee access to the first vaccines. As such, this program essentially eliminated issues with employee access which have been acknowledged as barriers to vaccination [32]. Nevertheless, concern over continued hesitancy and refusal in this group remains significant particularly in light of SARS-CoV-2 variant surges.

We formed an interdisciplinary team of health professionals and designed a study with a two-fold objective. First, we assessed levels of vaccine uptake categorized as acceptance, refusal or hesitancy in RUHS employees using an anonymous internet-based cross-sectional survey with direct employee solicitation. Second, we sought to understand determinants of decisions to vaccinate and of vaccine hesitancy and refusal. We focused on potential factors for which educational and other interventions could be targeted. This research paper offers results of a survey of healthcare workers fielded after the emergency use authorization (EUA) and use of the Pfizer, Moderna and Johnson & Johnson vaccines in the US.

## 2. Materials and Methods

### 2.1 Study Design and Population

Beginning in November 2020, a collaboration was established between the Comparative Effectiveness and Clinical Outcomes Research Center (CECORC) at Riverside University Health System (RUHS) and Claremont Graduate University (CGU). RUHS is an integrated health network in Riverside County, California that includes a 439-bed county Medical Center, 10 federally qualified health centers, several primary and specialty clinics, and the departments of Behavioral and Public Health. RUHS is a safety net California county health system which serves the over 2.3 million residents of Riverside County.

A cross-sectional survey to assess vaccine hesitancy among RUHS Medical Center employees was developed using survey information from previously published surveys of US and Canadian adults [19-22]. The RUHS-CECORC/CGU team met regularly to review and revise the survey and to plan for its administration. The finalized survey instrument was adapted for administration via Survey Monkey and took about 10 minutes to complete (available in Supplemental Materials). All responses and demographic data were collected from the survey participants directly and we did not use any hospital, medical or employee records.

RUHS Medical Center employees were eligible and invited to participate in the survey by an initial email followed by three subsequent reminder emails. RUHS-CECORC staff and volunteers distributed recruitment flyers in person at the medical center three mornings a week as workers entered their place of work. A total of 2,983 employees were eventually provided the survey. All respondents consented to participate in the survey by clicking “next” after introductory text and instructions on how to complete the survey.

The study was reviewed by the RUHS Institutional Review Board and classified as exempt as all responses were collected in a de-identified manner.

### 2.2 Survey Development and Measures

#### 2.1.1. Demographic and health-related characteristics

Questions on demographic characteristics used common US Census formats for response categories. Information on current job status at RUHS was self-reported. Respondents were asked if they had ever been told by a doctor or other health care provider if they had one or more underlying health conditions which would put them at higher risk for severe COVID-19 including diabetes, hypertension, asthma, serious heart conditions, chronic lung disease, chronic kidney disease, liver disease or a weakened immune system [14,15]. Individual conditions were summed to calculate total number of underlying health conditions/comorbidities. We used self-reported height and weight to calculate body mass index (BMI) and categorize participants into underweight/normal, overweight or obese.

#### 2.1.2. Experience with COVID-19

Our survey assessed employees’ perception of their exposure to COVID-19 on a weekly basis (no direct exposure, minimal, moderate or high exposure). Items asked whether the respondent or anyone they knew had ever tested positive for COVID-19. Given the relationship to the person, participants were asked to describe the severity of their symptoms [No Symptoms, Mild (symptoms but no shortness of breath), Moderate (visited doctor but no hospital stay), Severe/Critical (hospital stay), Death]. The impact of the pandemic on the respondent’s employment/income, mental and physical health, and ability to carry out normal activities was evaluated using a Likert scale with response options “severely decreased”, “decreased”, “no effect”, “improved”, “don’t know”.

Two sets of items assessed knowledge of COVID-19. Each correct response to a question which included six true/false items about COVID-19 disease was summed to create a disease knowledge scale (possible range 0-6). A second question asked respondents to identify common symptoms of COVID-19 from among 14 presented; each of ten correctly selected items were summed to create a symptom knowledge scale (possible range 0-10). For both scales, a higher score indicated greater knowledge.

#### 2.1.3. COVID-19 vaccine acceptance groups

Based on responses to three items about intent to receive a COVID-19 vaccination, we defined three groups (vaccine acceptors, hesitant, refusers) as follows. Respondents who reported having been vaccinated against COVID-19 (either fully or partially) or who planned to be vaccinated were categorized as vaccine acceptors. Those who reported not currently being vaccinated and were uncertain whether they would be vaccinated when an opportunity arises either now or at a future date were categorized as vaccine hesitant. Respondents who reported not currently being vaccinated, did not plan to be vaccinated when an opportunity arises, and would not consider vaccination at a later date were categorized as vaccine refusers.

#### 2.1.4. Vaccine hesitancy

To measure vaccine hesitancy, we included an adapted [22] version of the validated WHO SAGE working group vaccine hesitancy scale [2] for use in adults and implemented among health system workers [22]. The vaccine hesitancy scale is constructed using responses to eleven items which asked participants to rate their opinion on a Likert scale with response options “strongly disagree”, “disagree”, “neutral”, “agree”, “strongly agree”. For three items, a “strongly agree” response indicated a higher level of vaccine hesitancy. Other items were reverse-coded so as for their interpretation to be consistent. Scores on the scale ranged from 11-55 with higher scores indicating greater vaccine hesitancy.

#### 2.1.5. Determinants of vaccination

Depending on responses to the three items querying COVID-19 vaccination intention, participants were directed in the survey to a slightly different version of questions asking them about influences or potential influences of their decision to be vaccinated. Participants were presented with seventeen different determinants ranging from contextual influences (i.e., historical, socio-cultural, political, economic or health system/institutional factors), to individual and group influences (i.e., personal perception or social/peer environment), to vaccine-specific issues (i.e., directly related to COVID-19 vaccination). Determinants related to financial incentives were formulated based on prior studies [33-36]. All other items were modeled after Reiter et al. 2020 [20]; Pogue et al. 2020 [19]; Taylor et al. 2020 [21]. Respondents were asked to rank the level of influence on their decision to be vaccinated using a Likert Scale with response options “definitely would not”, “probably would not”, “not sure”, “probably would”, “definitely would”.

#### 2.1.6. Other items

We believed it likely that individuals who did not get a flu vaccine or have their children vaccinated [37,38] were also more likely to be vaccine refusers. Thus, questions about annual influenza vaccination were included for comparison to determine whether respondents took the flu vaccine as recommended (yes, no, skip some years). Respondents were asked to rank in order of importance a series of reasons understood to motivate flu vaccination including the safety and effectiveness of the vaccines; allergies to the vaccine; desire not to infect co-workers, patients or family members. In addition, opinions about the importance of childhood vaccinations were collected among participants with children.

### 2.3 Reliability of survey

Constructed scales for knowledge of COVID-19 in the final survey were assessed for intra-rater reliability using test-retest data from five non-survey participants to calculate intraclass correlations (ICCs) of averaged scale values for each rater. The ICC for the COVID-19 symptom knowledge scale was very good (ICC = 0.87) and for the COVID-19 disease knowledge scale was excellent (ICC > 0.99).

We examined the internal consistency of the vaccine hesitancy scale and the determinants of vaccination items by calculating Cronbach’s alpha among all respondents who completed the survey. Both the vaccine hesitancy scale and the determinants items demonstrated excellent internal consistency (standardized alpha = 0.92 and 0.93, respectively).

### 2.4 Statistical Analysis

Descriptive statistics (means, frequencies) for survey participants were summarized overall and compared between groups (vaccine acceptors, hesitant, refusers) using chi-square tests, Fisher’s exact test for categorical and t-tests or ANOVAs for continuous variables. Statistical tests were two-tailed.

We used general linear regression models to examine factors that were associated with vaccine hesitancy; in these models, the score on the vaccine hesitancy scale was the dependent outcome variable. Estimated β coefficients and standard errors (SE) of β expressed the average point difference in the vaccine hesitancy scale associated with a given variable compared to the reference group of that variable. β coefficients > 1 indicated greater hesitancy for the group compared with the reference; β coefficients < 1 indicated less hesitancy. We used multinomial logistic regression to identify predictors of COVID-19 vaccine acceptance comparing refusers and (separately) hesitant employees to acceptors. Estimated odds ratios (ORs) and 95% confidence intervals (CIs) expressed the increased or decreased likelihood associated with a given variable of being vaccine hesitant or a vaccine refuser compared with being an acceptor. Some variables were re-categorized due to small numbers of responders to that question. In model building approaches, all variables with global p < 0.10 in univariate analyses or for which the literature supported a relationship with vaccine hesitancy [6,8] were entered into a preliminary multivariable model. We then used stepwise procedures to retain variables with p < 0.05 in final multivariable linear and logistic models.

Responses to each of the seventeen different determinants influencing participants’ decision to be vaccinated were compared between the three groups of participants (vaccine acceptors, hesitant, refusers) using Kruskal-Wallis tests with Bonferroni adjustment to account for multiple testing (with the adjusted α set at 0.003). Effects of the COVID-19 pandemic on employment, income and health were similarly compared between the three groups without Bonferroni adjustment. Analyses used SAS version 9.4 (SAS Institute Inc., Cary, NC, USA) or RStudio version 1.3 1093 (2009-2020 RStudio, PBC, Boston, MA, USA).

## 3. Results

The survey was administered from March 15 – April 26, 2021 to 2,983 RUHS Medical Center employees; 791 surveys were returned for a 27% response rate. After excluding 2 records because most responses were blank or in one case, appeared to be fictitious, 789 remained. Respondents were predominantly female (79.2%), between the ages of 30-64 (83.7%), non-Hispanic white (37.7%) or Hispanic (36.8%) and had a self-reported education level of a college degree or higher (59.7%). Of those responding to the survey, 755 (95.6%) answered questions enabling a categorization into groups of vaccine acceptors, hesitant, or refusers. Vaccine hesitant and refusers were more likely to be women and to have an annual household income of less than $50,000. Refusers were more likely to be in the 30-49-year age range. A greater proportion of vaccine hesitant had less than a college degree, and both hesitant and refusers had lower proportions with a college degree or higher (Table 1).

**Table 1.**
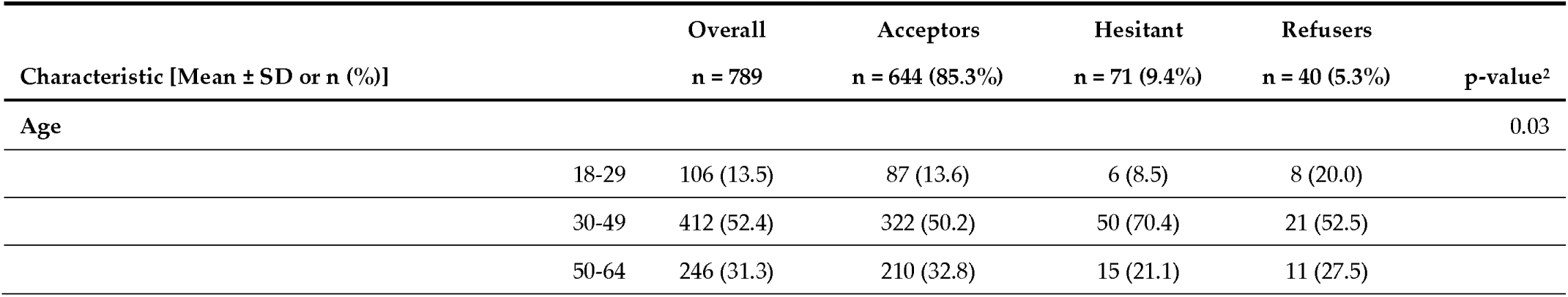
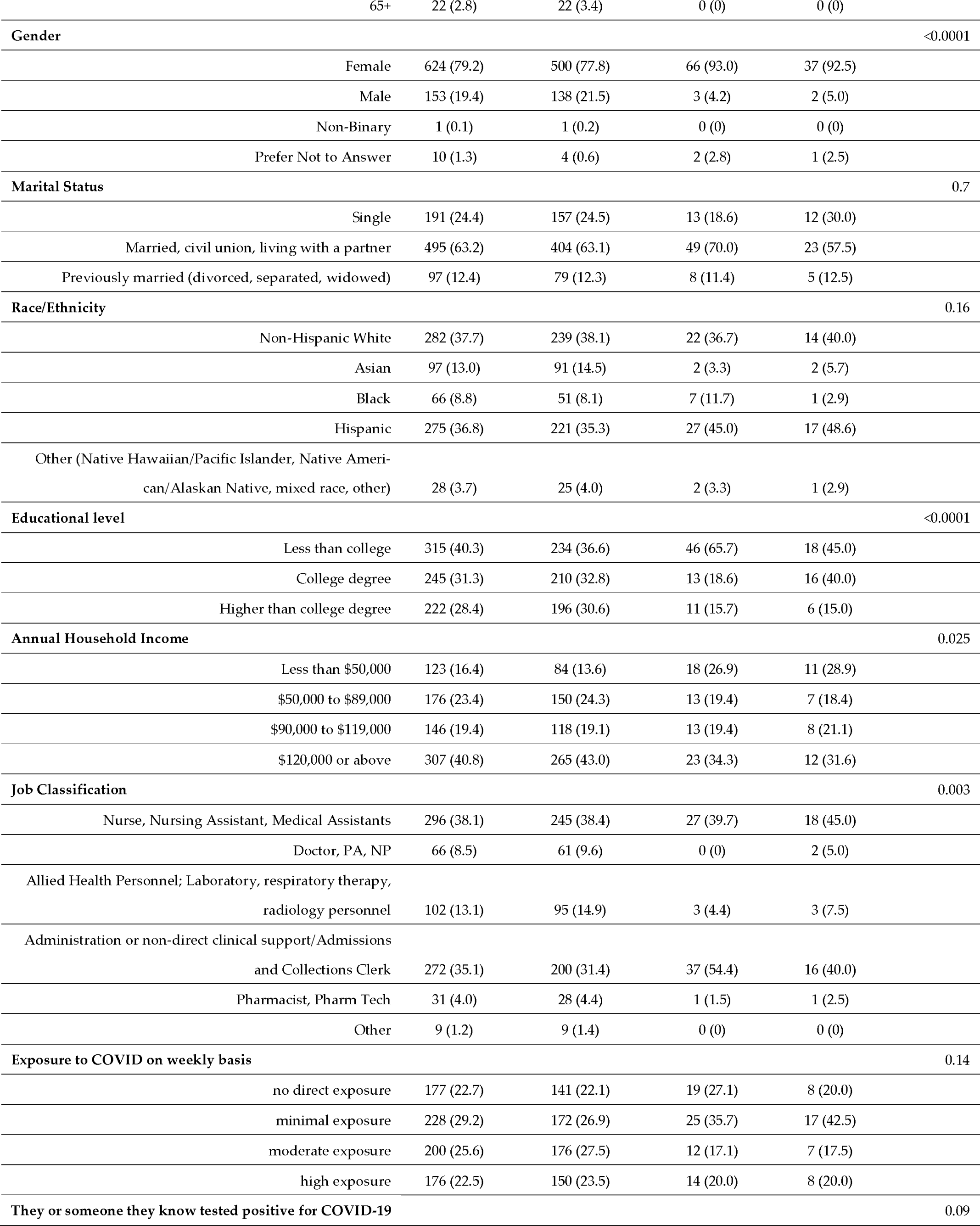
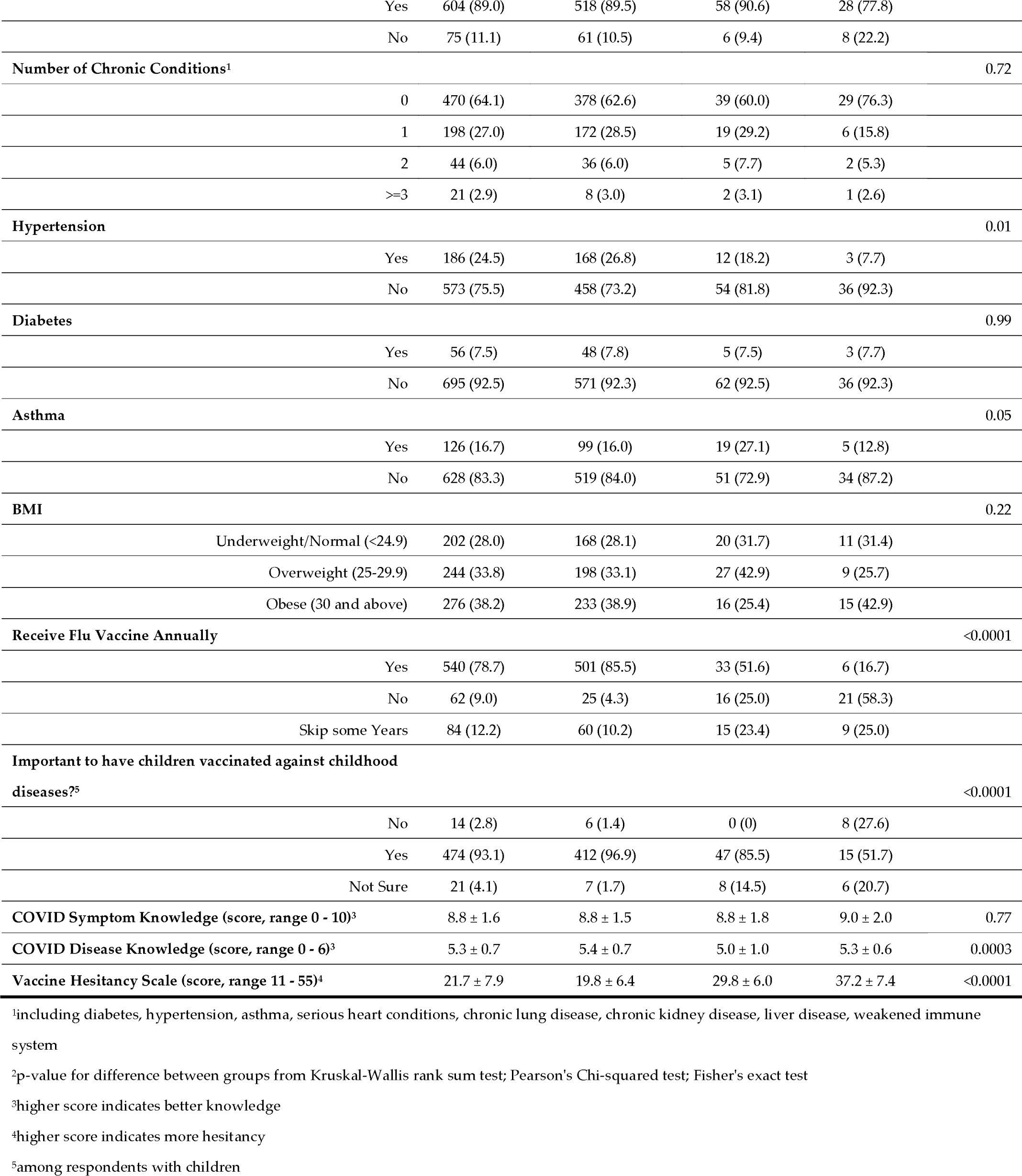
Characteristics of RUHS Medical Center Employees (n=789) who Participated in COVID-19 Vaccine Survey, by Vaccine Acceptance Status (Acceptors, Hesitant, Refusers)

| Characteristic [Mean ± SD or n (%)] | Overall<br>n = 789 | Acceptors<br>n = 644 (85.3%) | Hesitant<br>n = 71 (9.4%) | Refusers<br>n = 40 (5.3%) | p-value<sup>2</sup> |
| --- | --- | --- | --- | --- | --- |
| Age |  |  |  |  | 0.03 |
| 18-29 | 106 (13.5) | 87 (13.6) | 6 (8.5) | 8 (20.0) |  |
| 30-49 | 412 (52.4) | 322 (50.2) | 50 (70.4) | 21 (52.5) |  |
| 50-64 | 246 (31.3) | 210 (32.8) | 15 (21.1) | 11 (27.5) |  |

|  |  |  |  |  |  |  |
| --- | --- | --- | --- | --- | --- | --- |
|  | 65+ | 22 (2.8) | 22 (3.4) | 0 (0) | 0 (0) |  |
| **Gender** |  |  |  |  |  | <0.0001 |
|  | Female | 624 (79.2) | 500 (77.8) | 66 (93.0) | 37 (92.5) |  |
|  | Male | 153 (19.4) | 138 (21.5) | 3 (4.2) | 2 (5.0) |  |
|  | Non-Binary | 1 (0.1) | 1 (0.2) | 0 (0) | 0 (0) |  |
|  | Prefer Not to Answer | 10 (1.3) | 4 (0.6) | 2 (2.8) | 1 (2.5) |  |
| **Marital Status** |  |  |  |  |  | 0.7 |
|  | Single | 191 (24.4) | 157 (24.5) | 13 (18.6) | 12 (30.0) |  |
|  | Married, civil union, living with a partner | 495 (63.2) | 404 (63.1) | 49 (70.0) | 23 (57.5) |  |
|  | Previously married (divorced, separated, widowed) | 97 (12.4) | 79 (12.3) | 8 (11.4) | 5 (12.5) |  |
| **Race/Ethnicity** |  |  |  |  |  | 0.16 |
|  | Non-Hispanic White | 282 (37.7) | 239 (38.1) | 22 (36.7) | 14 (40.0) |  |
|  | Asian | 97 (13.0) | 91 (14.5) | 2 (3.3) | 2 (5.7) |  |
|  | Black | 66 (8.8) | 51 (8.1) | 7 (11.7) | 1 (2.9) |  |
|  | Hispanic | 275 (36.8) | 221 (35.3) | 27 (45.0) | 17 (48.6) |  |
|  | Other (Native Hawaiian/Pacific Islander, Native American/Alaskan Native, mixed race, other) | 28 (3.7) | 25 (4.0) | 2 (3.3) | 1 (2.9) |  |
| **Educational level** |  |  |  |  |  | <0.0001 |
|  | Less than college | 315 (40.3) | 234 (36.6) | 46 (65.7) | 18 (45.0) |  |
|  | College degree | 245 (31.3) | 210 (32.8) | 13 (18.6) | 16 (40.0) |  |
|  | Higher than college degree | 222 (28.4) | 196 (30.6) | 11 (15.7) | 6 (15.0) |  |
| **Annual Household Income** |  |  |  |  |  | 0.025 |
| | Less than \$50,000 | 123 (16.4) | 84 (13.6) | 18 (26.9) | 11 (28.9) | |
| | \$50,000 to \$89,000 | 176 (23.4) | 150 (24.3) | 13 (19.4) | 7 (18.4) | |
| | \$90,000 to \$119,000 | 146 (19.4) | 118 (19.1) | 13 (19.4) | 8 (21.1) | |
| | \$120,000 or above | 307 (40.8) | 265 (43.0) | 23 (34.3) | 12 (31.6) | |
| **Job Classification** |  |  |  |  |  | 0.003 |
|  | Nurse, Nursing Assistant, Medical Assistants | 296 (38.1) | 245 (38.4) | 27 (39.7) | 18 (45.0) |  |
|  | Doctor, PA, NP | 66 (8.5) | 61 (9.6) | 0 (0) | 2 (5.0) |  |
|  | Allied Health Personnel; Laboratory, respiratory therapy, radiology personnel | 102 (13.1) | 95 (14.9) | 3 (4.4) | 3 (7.5) |  |
|  | Administration or non-direct clinical support/Admissions and Collections Clerk | 272 (35.1) | 200 (31.4) | 37 (54.4) | 16 (40.0) |  |
|  | Pharmacist, Pharm Tech | 31 (4.0) | 28 (4.4) | 1 (1.5) | 1 (2.5) |  |
|  | Other | 9 (1.2) | 9 (1.4) | 0 (0) | 0 (0) |  |
| **Exposure to COVID on weekly basis** |  |  |  |  |  | 0.14 |
|  | no direct exposure | 177 (22.7) | 141 (22.1) | 19 (27.1) | 8 (20.0) |  |
|  | minimal exposure | 228 (29.2) | 172 (26.9) | 25 (35.7) | 17 (42.5) |  |
|  | moderate exposure | 200 (25.6) | 176 (27.5) | 12 (17.1) | 7 (17.5) |  |
|  | high exposure | 176 (22.5) | 150 (23.5) | 14 (20.0) | 8 (20.0) |  |
| **They or someone they know tested positive for COVID-19** |  |  |  |  |  | 0.09 |
|  | Yes | 604 (89.0) | 518 (89.5) | 58 (90.6) | 28 (77.8) |  |
|  | No | 75 (11.1) | 61 (10.5) | 6 (9.4) | 8 (22.2) |  |
| **Number of Chronic Conditions<sup>1</sup>** |  |  |  |  |  | 0.72 |
|  | 0 | 470 (64.1) | 378 (62.6) | 39 (60.0) | 29 (76.3) |  |
|  | 1 | 198 (27.0) | 172 (28.5) | 19 (29.2) | 6 (15.8) |  |
|  | 2 | 44 (6.0) | 36 (6.0) | 5 (7.7) | 2 (5.3) |  |
|  | ≥3 | 21 (2.9) | 8 (3.0) | 2 (3.1) | 1 (2.6) |  |
| **Hypertension** |  |  |  |  |  | 0.01 |
|  | Yes | 186 (24.5) | 168 (26.8) | 12 (18.2) | 3 (7.7) |  |
|  | No | 573 (75.5) | 458 (73.2) | 54 (81.8) | 36 (92.3) |  |
| **Diabetes** |  |  |  |  |  | 0.99 |
|  | Yes | 56 (7.5) | 48 (7.8) | 5 (7.5) | 3 (7.7) |  |
|  | No | 695 (92.5) | 571 (92.3) | 62 (92.5) | 36 (92.3) |  |
| **Asthma** |  |  |  |  |  | 0.05 |
|  | Yes | 126 (16.7) | 99 (16.0) | 19 (27.1) | 5 (12.8) |  |
|  | No | 628 (83.3) | 519 (84.0) | 51 (72.9) | 34 (87.2) |  |
| **BMI** |  |  |  |  |  | 0.22 |
|  | Underweight/Normal (<24.9) | 202 (28.0) | 168 (28.1) | 20 (31.7) | 11 (31.4) |  |
|  | Overweight (25-29.9) | 244 (33.8) | 198 (33.1) | 27 (42.9) | 9 (25.7) |  |
|  | Obese (30 and above) | 276 (38.2) | 233 (38.9) | 16 (25.4) | 15 (42.9) |  |
| **Receive Flu Vaccine Annually** |  |  |  |  |  | <0.0001 |
|  | Yes | 540 (78.7) | 501 (85.5) | 33 (51.6) | 6 (16.7) |  |
|  | No | 62 (9.0) | 25 (4.3) | 16 (25.0) | 21 (58.3) |  |
|  | Skip some Years | 84 (12.2) | 60 (10.2) | 15 (23.4) | 9 (25.0) |  |
| **Important to have children vaccinated against childhood diseases<sup>25</sup>** |  |  |  |  |  | <0.0001 |
|  | No | 14 (2.8) | 6 (1.4) | 0 (0) | 8 (27.6) |  |
|  | Yes | 474 (93.1) | 412 (96.9) | 47 (85.5) | 15 (51.7) |  |
|  | Not Sure | 21 (4.1) | 7 (1.7) | 8 (14.5) | 6 (20.7) |  |
| **COVID Symptom Knowledge (score, range 0 - 10)<sup>3</sup>** |  | 8.8 ± 1.6 | 8.8 ± 1.5 | 8.8 ± 1.8 | 9.0 ± 2.0 | 0.77 |
| **COVID Disease Knowledge (score, range 0 - 6)<sup>3</sup>** |  | 5.3 ± 0.7 | 5.4 ± 0.7 | 5.0 ± 1.0 | 5.3 ± 0.6 | 0.0003 |
| **Vaccine Hesitancy Scale (score, range 11 - 55)<sup>4</sup>** |  | 21.7 ± 7.9 | 19.8 ± 6.4 | 29.8 ± 6.0 | 37.2 ± 7.4 | <0.0001 |

<sup>1</sup>including diabetes, hypertension, asthma, serious heart conditions, chronic lung disease, chronic kidney disease, liver disease, weakened immune system
<sup>2</sup>p-value for difference between groups from Kruskal-Wallis rank sum test; Pearson's Chi-squared test; Fisher's exact test
<sup>3</sup>higher score indicates better knowledge
<sup>4</sup>higher score indicates more hesitancy
<sup>5</sup>among respondents with children

Job classifications of survey respondents were generally reflective of the overall makeup of medical center employees of 45% nurses, 5% physicians, 19% ancillary and 30% non-medical personnel, based on RUHS human resources data. Higher proportions of nurses, nursing assistants and medical assistants were among the vaccine refusers and higher proportions of administrative, non-clinical staff were among the vaccine hesitant. Physicians and allied health personnel were more likely to be in the vaccine acceptor group. Respondents were approximately evenly distributed by level of exposure to COVID-19 on a weekly basis (Table 1). Eighty-nine percent either had themselves or knew someone who had tested positive for COVID-19.

Twenty-seven percent of respondents reported having one chronic condition, with hypertension being the most common (24.5%), followed by asthma (16.7%) and diabetes (7.5%), and 72% had BMIs in the overweight or obese range, based on self-reported height and weight. Vaccine hesitant and refusers were less likely to have hypertension than vaccine acceptors (p = 0.01), but individuals who were vaccine hesitant were more likely to have asthma (p = 0.05).

Seventy-nine percent of respondents received the flu vaccine each year, which differed markedly between groups, with 85% of acceptors, 52% of hesitant and 17% of refusers reporting flu vaccination annually. Among respondents with children, 93% overall reported believing in the importance of having their children vaccinated against childhood diseases. This differed significantly between groups with nearly all of vaccine acceptors (97%), 86% of vaccine hesitant, yet just over half (52%) of vaccine refusers reporting affirmatively.

Respondents as a group were knowledgeable about both COVID-19 symptoms and disease, with average scores on corresponding scales near the upper bound of the total possible. Vaccine hesitant had lower knowledge of COVID-19 disease compared with acceptors and refusers (p=0.0003).

Scores on the vaccine hesitancy scale tracked, as anticipated, based on the three categories of respondents. Overall, respondents’ scores on the vaccine hesitancy scale were 21.7 out of 55, with scale scores differentiating well between the three groups (acceptors, hesitant, refusers) (Table 1). Scores were lowest among vaccine acceptors, while vaccine refusers had the highest scores, indicating the greatest hesitancy. An average of 10 points separated the vaccine hesitant from the vaccine acceptors, and an average of 18 separated the vaccine refusers from the vaccine acceptors (p <0.0001).

Eleven of the 23 characteristics were individually associated with vaccine hesitancy as assessed with the vaccine hesitancy scale: gender, race/ethnicity, marital status, educational level, annual household income, job classification, weekly exposure to COVID-19, annual flu vaccination, importance of childhood vaccinations and COVID-19 disease knowledge (Table 2). Interestingly, several factors were not correlated with hesitancy including age, testing positive or knowing someone who tested positive for COVID-19, BMI, or knowledge of COVID-19 symptoms. Despite well-characterized associations with medical complications and poor outcomes [39], no specific chronic condition nor the total number of chronic conditions were associated with vaccine hesitancy in our healthcare worker population. In multivariable models, race/ethnicity, marital status, job classification, immunocompromised status, annual flu vaccination and importance of childhood vaccinations were significant independent predictors of vaccine hesitancy. Asian respondents were more likely to be hesitant than non-Hispanic whites (β = 1.63; p = 0.06) and immunocompromised persons were more hesitant than those who were not (β = 2.36; p = 0.03). Compared to doctors, physician assistants and nurse practitioners, all other job classifications had higher vaccine hesitancy scores.

**Table 2.**
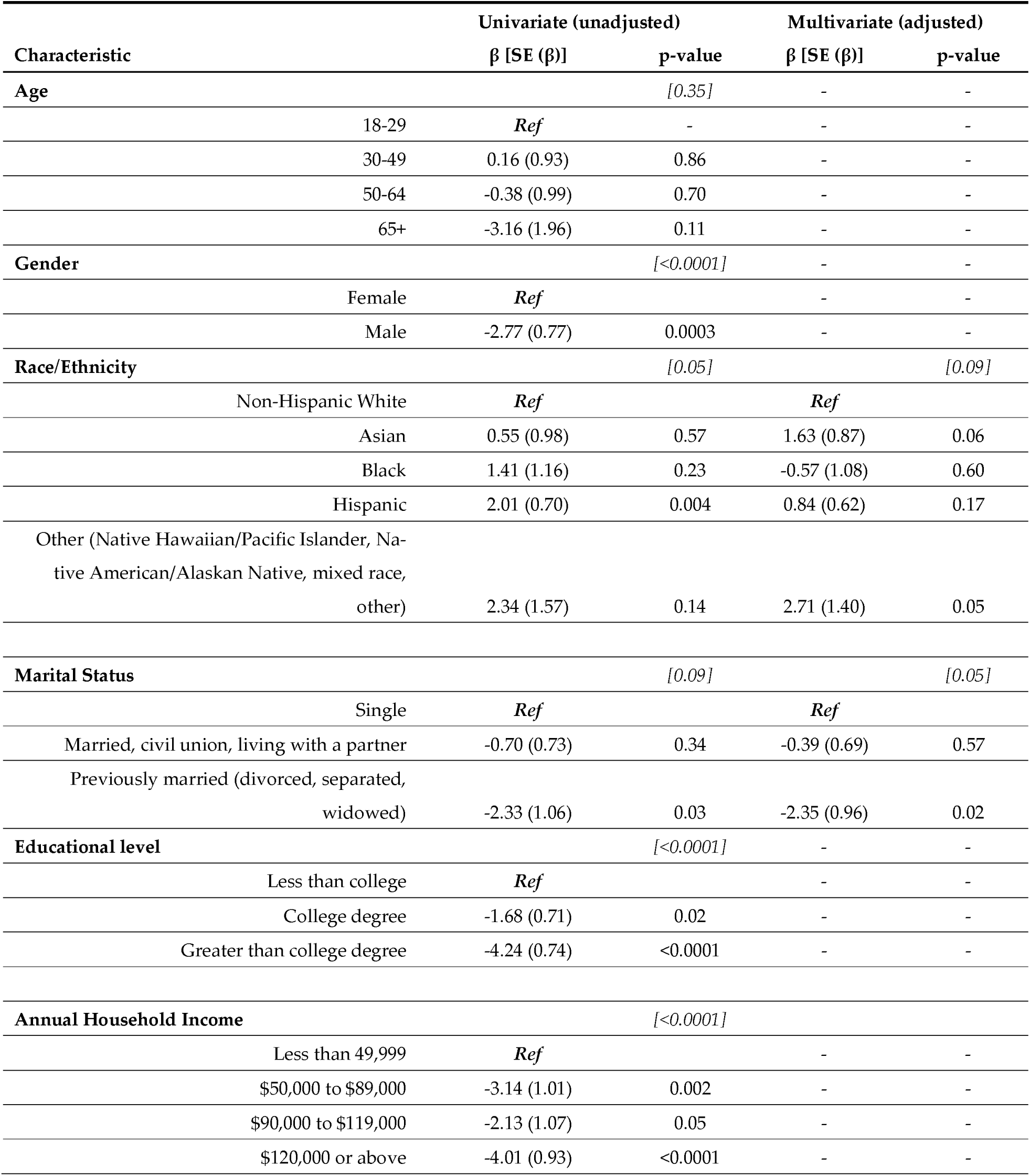
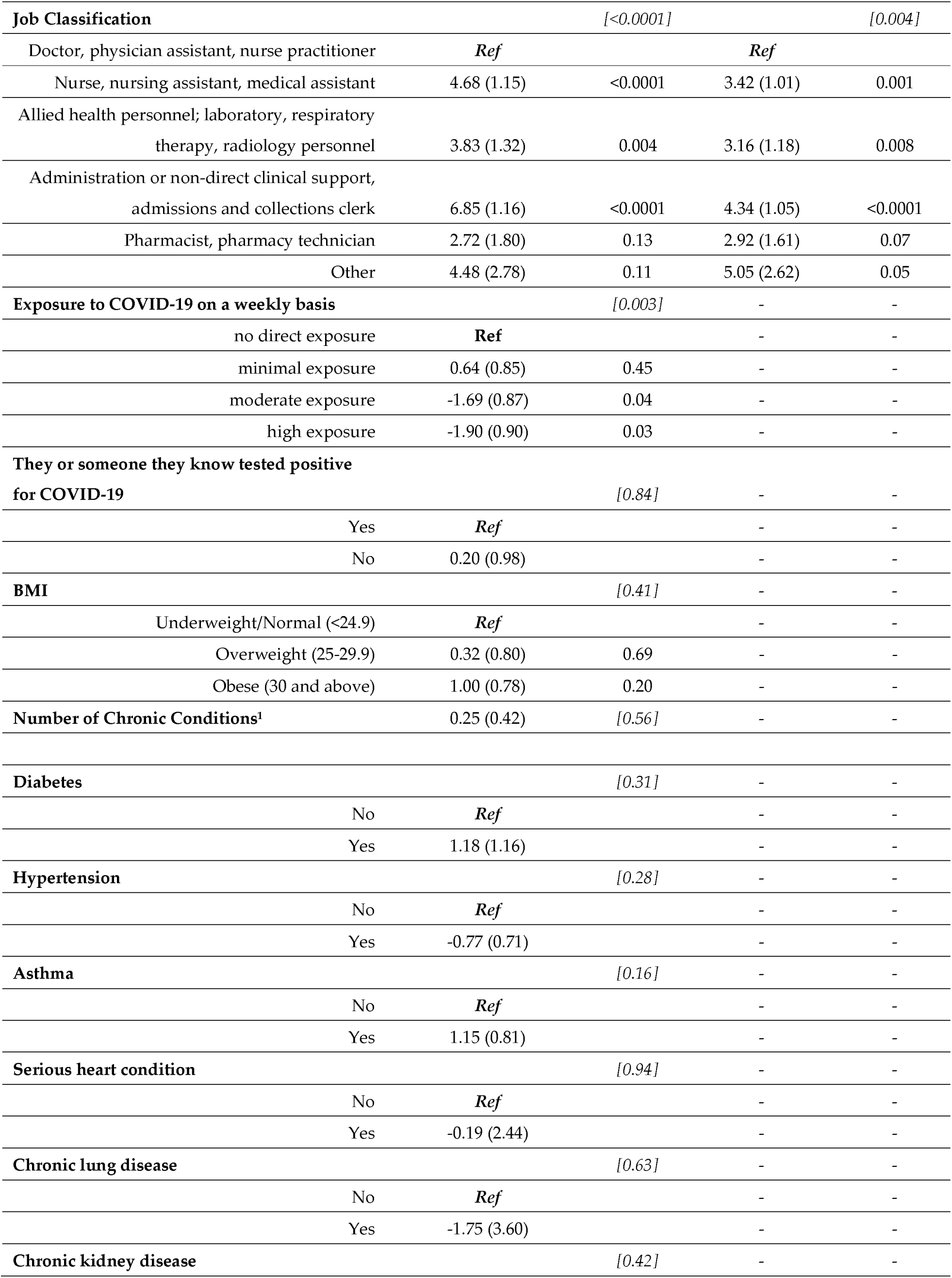
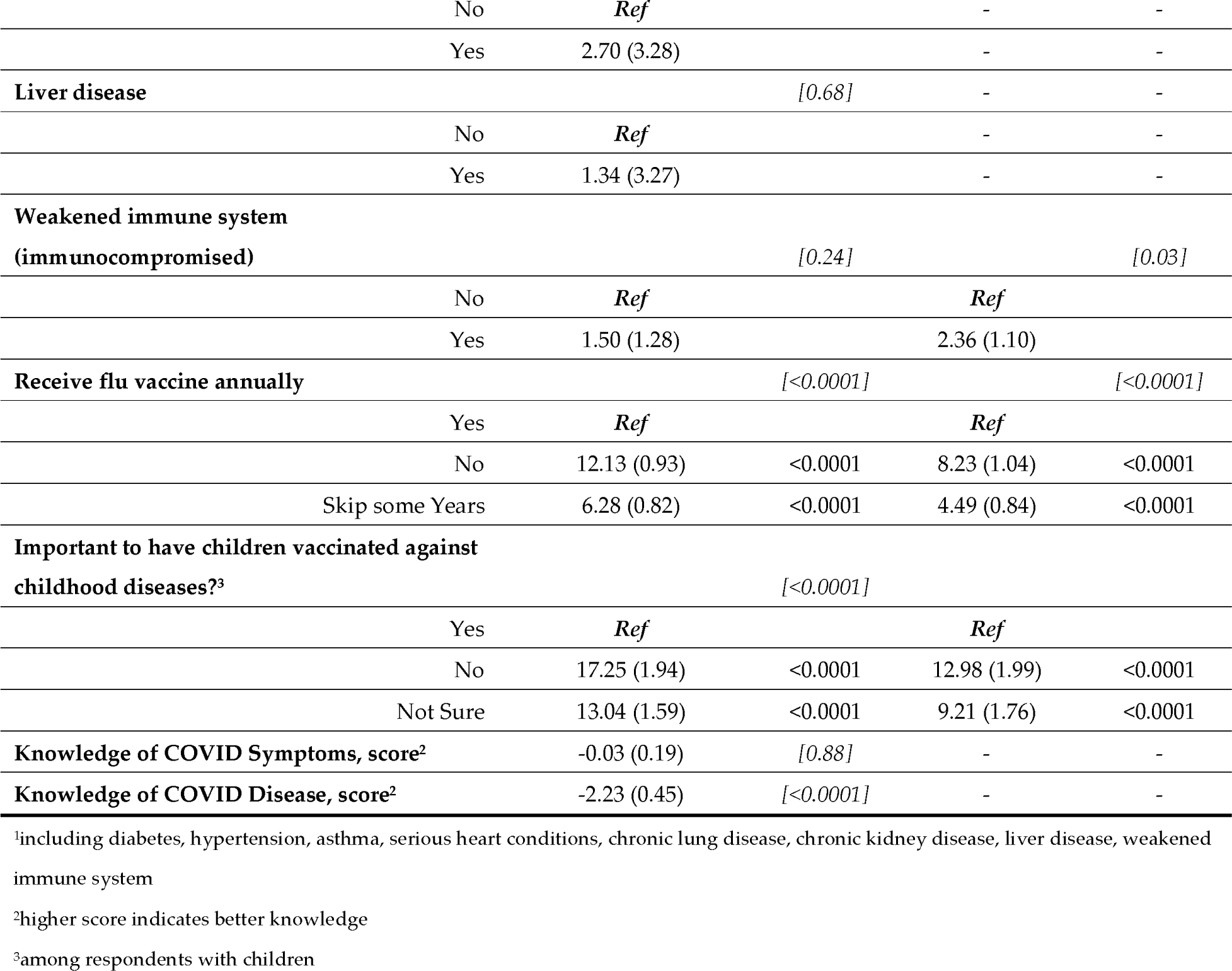
Univariate and multivariate associations (β [SE (β)]; p-value) between potential predictors of the vaccine hesitancy scale from general linear regression models.

Acceptance of the flu vaccine was predictive of vaccine hesitancy, as employees who reported regularly receiving the flu vaccine scored an average of 8.23 points higher on the vaccine hesitancy scale (p < 0.0001) compared to those that did not, and persons who skipped some years scored an average of 4.49 points higher (p < 0.0001).

Employees who did not believe it was important to have their children vaccinated against childhood diseases scored nearly 13 points higher on the vaccine hesitancy scale compared to those who did have this belief (p < 0.0001), and those who were not sure about childhood vaccinations scored over 9 points higher (p < 0.0001). Respondents who were previously married were less likely to be hesitant than those who were currently single (β = -2.35; p = 0.02)

Several characteristics were individually associated with vaccine acceptance status including age, gender, educational level, job classification, BMI, testing positive or knowing someone who tested positive for COVID-19, hypertension, asthma, COVID-19 disease knowledge, annual flu vaccination, and importance of childhood vaccinations (Table 3). ORs estimated for the latter two had very wide confidence intervals due to the small number of observations in cells despite re-categorizing; as such, these variables were not further pursued in models building approaches. In multivariable models, gender, educational level, job classification and BMI were significantly independently predictive of vaccine acceptance, hesitancy and refusal. Compared with women, men were significantly less likely to be vaccine hesitant (OR = 0.12; 95% CI = 0.02, 0.91). Persons with a college degree or higher were 55% less likely to be vaccine hesitant compared to those with less than a college degree (OR = 0.44; 95% CI = 0.22, 0.89) and employees with a BMI classified as obese were 65% less likely to be vaccine hesitant compared to those with a BMI in the normal/underweight range (OR = 0.35; 95% CI = 0.14, 0.90). Non-clinical staff were more than twice as likely to be vaccine hesitant than clinical employees (OR = 2.31; 95% CI = 1.16, 4.59). Associations for these characteristics were apparent and in the same direction for vaccine refusers with the exception of BMI but did not achieve statistical significance likely because of the small numbers of refusers.

**Table 3.**
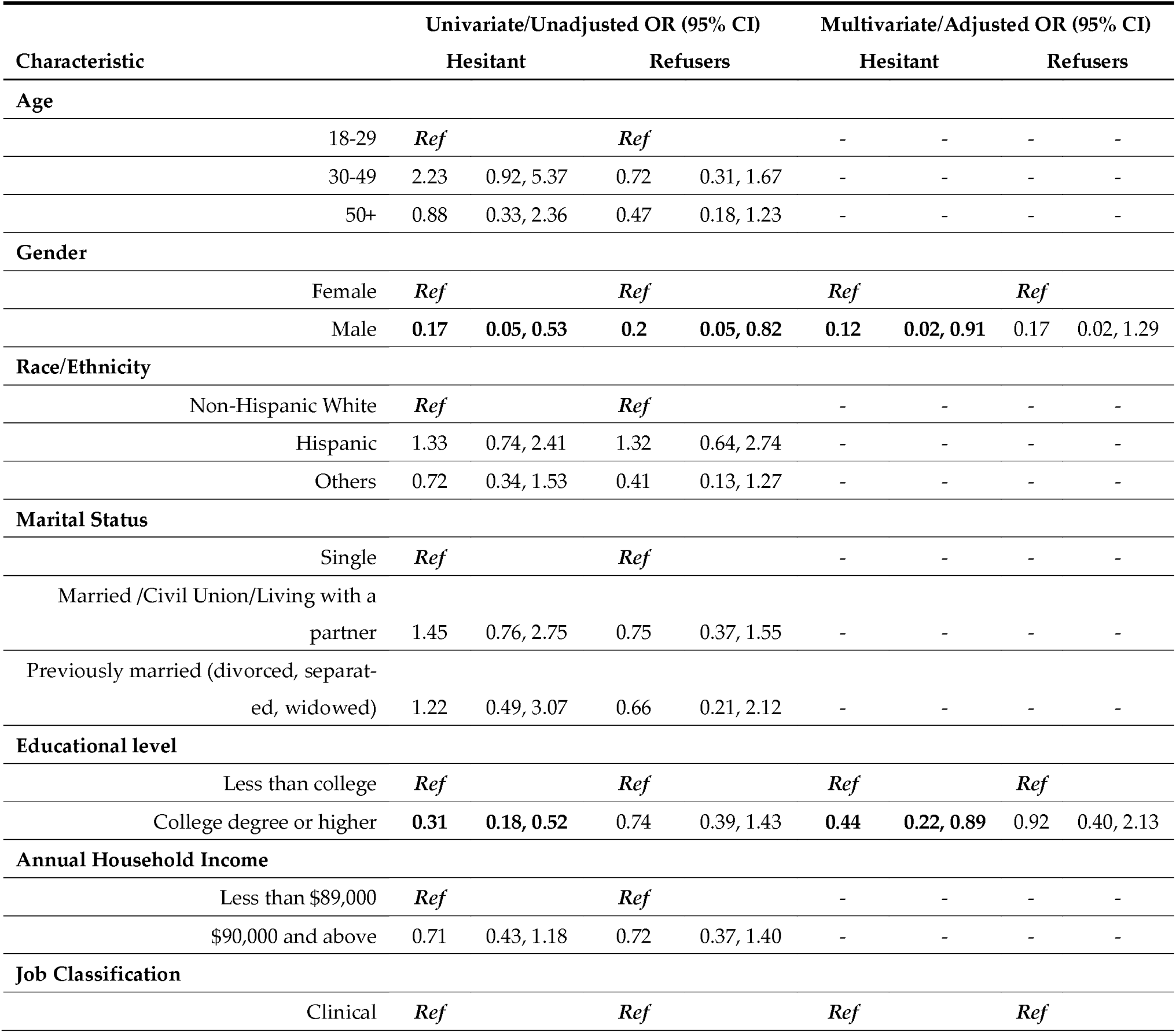
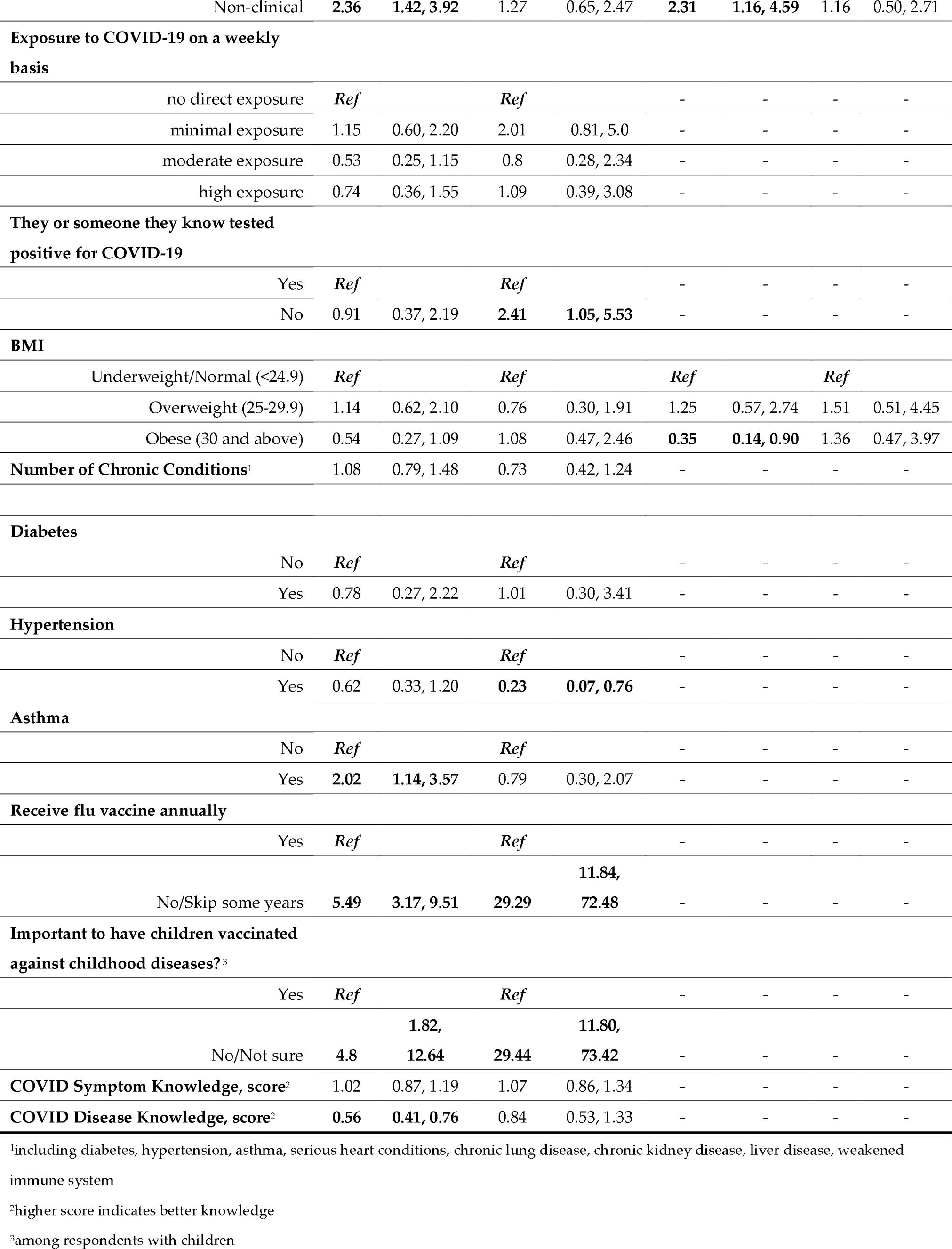
Univariate and multivariate associations (ORs, 95% CIs) between potential predictors of the vaccine acceptance, hesitancy or refusal from multinomial logistic regression models.

Employees at RUHS Medical Center who accepted COVID-19 vaccination reported several influences of their decision to be vaccinated, including protection of the vulnerable, encouragement from family members or colleagues, and advice from a health care worker. They also prioritized safety and efficacy of the vaccine and job or other requirements as determining factors. For those employees who accepted vaccination, financial incentives, raffles, social media and religious leaders were not motivating factors (Figure 1). Among RUHS employees who were hesitant, uncertainty was also prevalent in their reports of motivating factors influencing their indecision to accept or refuse vaccination, with many responses tending to concentrate along the middle of the Likert scale. Employees who refused vaccination, on the other hand, showed a very different pattern in their responses. In this group, all possible reasons were ranked as not impacting a potential decision to be vaccinated. Additionally, refusers indicated that financial incentives would not increase their likelihood of being vaccinated.

**Figure 1.**
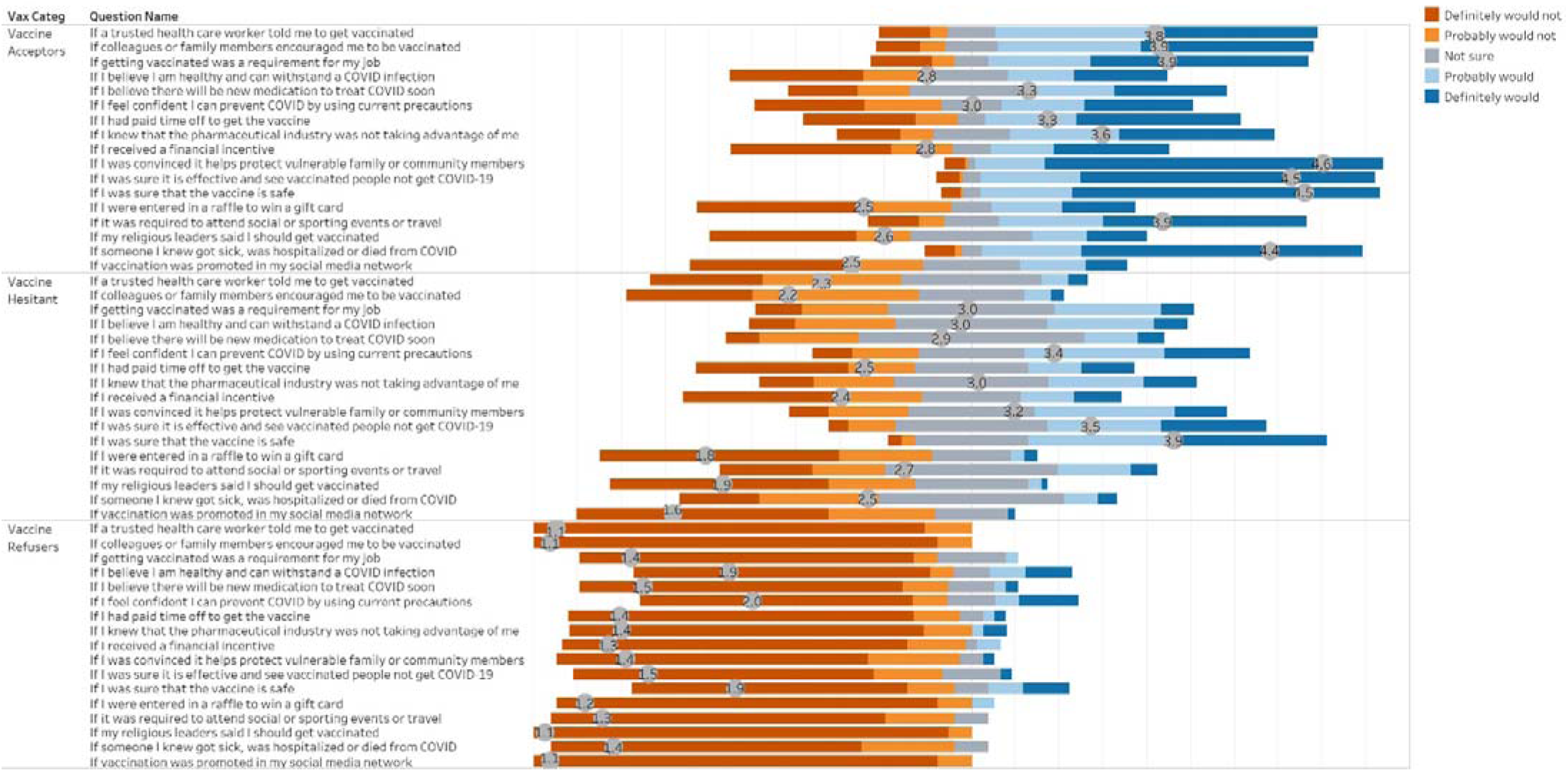
Influences of Decision for COVID-19 Vaccination with Average Score on Likert Scale for Each Response, by Group.

Despite a strong employee-support environment and job protection, respondents reported that the COVID-19 pandemic had affected their physical and mental health and well-being as well as their ability to carry out normal activities (Figure 2). Paradoxically, vaccine refusers reported their physical health was less affected than that reported by acceptors or hesitant. Both family income and employment were less impacted for all groups.

**Figure 2.**
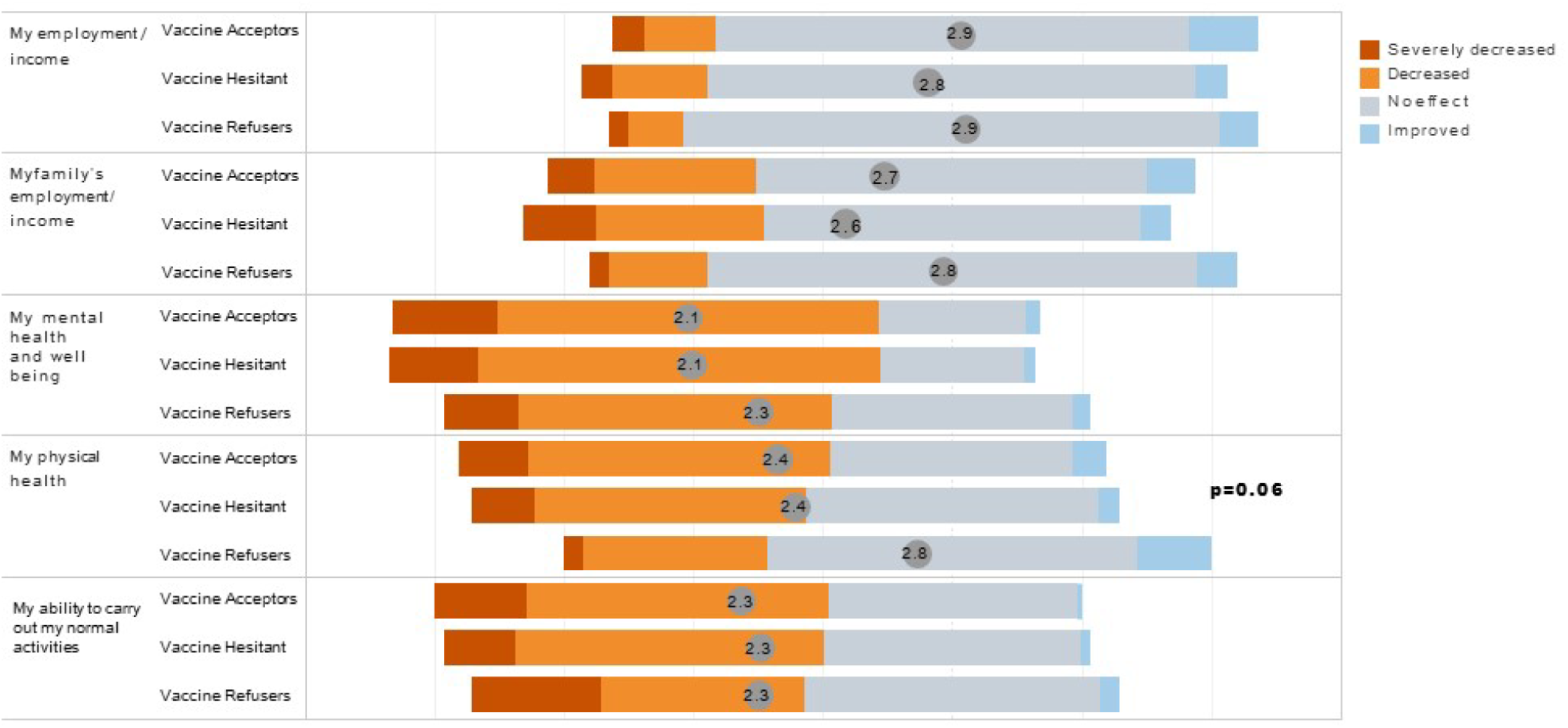
Reported effects of the COVID-19 pandemic on employment, income, health and normal activities, by group.

## 4. Discussion

In our comprehensive survey of employees of a large safety net county health system conducted between March and April 2021 when COVID-19 vaccinations were being administered in the US, we found that 9.4% overall were hesitant and 5.3% refused to be vaccinated, which is consistent with studies that have estimated 8-18% hesitancy [3]. These proportions contrast markedly from general population estimates of 20-30% resistant or refusing vaccination [13-16,23].

Similar to previous research among US healthcare workers [22,40-45], our data suggest that employees who hesitated or refused to be vaccinated against COVID-19 were more likely to be women, of younger ages, and to have lower levels of education and income. Also analogous to other studies that found less hesitancy among healthcare workers in direct patient care roles[22,42-45], we observed that employees with high or moderate weekly exposure to COVID-19 such as from working in the COVID ward or ICU in any capacity were less likely to be hesitant. Unlike some previous surveys [22,42-45] [46] we did not find lower hesitancy among Asian healthcare workers. One possible explanation for this difference is that the makeup of our study population within the Asian racial group is not comparable to those of previous research, which may be supported by another California-based study [23]. Our survey did not query the specific Asian designation i.e., Filipino, Chinese etc. nor did previous studies provide data to assess this.

One of our objectives was to potentially identify reasons underlying vaccine hesitancy and refusal in healthcare workers which could be targeted by interventions. As suggested by others [47], our findings provide the assessment of barriers to vaccination adoption in an organizational setting which can be used to identify evidence-based strategies to increase COVID-19 vaccine uptake. Based on responses from employees who were hesitant to be vaccinated, we suggest campaigns which focus on providing information and reassurances[48] regarding the safety and efficacy of COVID-19 vaccines. Efforts directed at building trust in vaccine manufacturers could also be useful as well as vaccine mandates as a condition of employment. We see an opportunity in educational programs given that employees who were vaccine hesitant had lower knowledge of COVID-19 disease, despite employment within a health system. Our data further suggest that messaging from colleagues or family members, within social media networks or by religious leaders is less likely to sway hesitant health system employees to be vaccinated. This finding can be supported by a similar study [41] which found that social media is not perceived a valuable source of vaccine information for healthcare workers given social media networks are platforms found to disseminate extensive vaccine misinformation[49]. The response pattern among those who refused to be vaccinated indicates that converting these persons will likely be more challenging as no one reason emerged as a potential candidate for a targeted intervention. Similar to previous research [42,45,46], acceptance of annual flu vaccination and the importance of childhood vaccination were significant predictors of COVID-19 vaccine hesitancy and refusal in our population. These likely reflect longer standing beliefs which may be more difficult to modify. Therefore, in order to address vaccine hesitancy and increase COVID-19 vaccine uptake, it will be important to examine beliefs of healthcare workers with pre-existing concerns about vaccines in general.

Response bias may be a consideration as participants in the survey did so voluntarily. We suspect that both vaccine hesitant and refusers are higher within our employees as these groups may not have responded to the survey since they may hold underlying concerns and suspicions related to the intention of the questions despite reassurance of anonymity. Additionally, these two groups may not have completed a survey because of perceived stigmas associated with or the social undesirability of their responses. On the other hand, those that have accepted vaccination may find little personal value in completing a survey that largely has no direct relevance to their decision. In sum, it is likely that vaccine hesitant and refusers are underestimated in our study, and those who did participate may not be representative of all RUHS health system employees.

It is important to contextualize the survey with the historical events occurring contemporaneously with when data were being collected in March and April 2021. Globally, countries marked the 1-year anniversary of the COVID-19 pandemic and the WHO released a report on the potential origin of the virus in China [2]. Debate about the origin of the SARS-CoV-2 virus continued to evolve and was driven by deep-seated political beliefs. In the US, three vaccines had EUA by the FDA, two had been in use since January 2021. The Biden administration announced the purchase of additional Johnson & Johnson vaccines in order to expand supply to have sufficient vaccines for all US adults by the end of May 2021. This preceded the pause in use of their vaccine over concerns of blood clotting. Pfizer & Modera mRNA vaccines were found to prevent infection not just illness, and both manufacturers began trials on children aged 6 months – 11 years. In California, vaccine eligibility was expanded to additional groups, and some “lockdown” measures were relaxed as some counties moved into less restrictive tiers. Changing public health recommendations as well as the scientific complexity related to the novel coronavirus and our understanding as to how the virus adapts impacted the perceived confidence of the general population for vaccination. Different from previous studies among healthcare workers, by the time our survey was administered, vaccination against COVID-19 was not a hypothetical. The majority of published US studies[22,24,42,44,45] that surveyed healthcare workers were during periods when vaccines were still under development; others coincided with early vaccination efforts of healthcare workers and nursing home residents[40,41,43,46]. Moreover, our Medical Center had an early and robust employee focused vaccination program at the time the survey was administered.

The findings of this study should be interpreted cautiously given that they are based on data from 71 employees whose survey responses indicated they were hesitant to be vaccinated and 40 whose responses indicated they refused vaccination. As with other cross-sectional surveys, we do not have longitudinal data to examine if positions about COVID-19 vaccination changed over time among our health system employee cohort. Thus, the survey responses reflect a “snapshot” of opinions at one point in time and should be contextualized as described above. We plan a follow-up survey of employees to examine change in vaccine acceptance. One major strength of our study over previous research is our multivariable modeling approach which identifies factors that are independent predictors of vaccine hesitancy and refusal. This is useful because individual factors are often correlated, eg., education, income, and job title, and may be reflective of a common underlying construct. An additional strength of our work is our implementation of two measures of vaccine hesitancy, one derived from survey items and a second from a validated vaccine hesitancy scale [2]. Furthermore, we demonstrated that essential components of our survey instrument were reliable.

## 5. Conclusions

Vaccine hesitancy including refusing vaccination is a major global concern and is not novel to the COVID-19 pandemic [4-6]. Continued attempts to reassure people of the efficacy and safety of vaccines and to accept vaccination for influenza and other seasonal virus infections has been an ongoing public health effort for decades [50]. This study demonstrates that healthcare workers are not immune to concerns related to vaccination. A troubling finding of our study is the effect of the pandemic on wellbeing and work performance within health system employees despite strong benefit support, salary protection, healthcare benefits and continued employment assurance. Implications of the reported short-term effects and potential for long-term consequences merit further investigation.

Unvaccinated persons are both victims and culprits of SARS-CoV-2. The notion that the virus would be completely controlled with high vaccination rates has been challenged by the emergence of more infectious variants like delta together with significant proportions of vaccine hesitant persons in populations. This further raises concerns that additional variants may challenge the immune protection now afforded by current vaccines. Therefore, it is important to understand the motivations and beliefs of those not accepting vaccinations and to develop interventions that may increase acceptance, particularly among healthcare workers who are in positions to influence others. We point to our observation that diverged from most previous work, of greater hesitancy among Asian health system employees as a reason why a “one-size fits all” approach will not suffice as others similarly advocate [51]. Future research surveys and other quantitative analyses should be accompanied by qualitative research aimed at discovering more information on why individuals have refused vaccinations and more importantly, what can be done about it. We recommend that focus groups of refusers and vaccine hesitant healthcare workers lead to a thoughtful and deep probing of reasons and a full discussion of strategies suggested by participants that could result in their being vaccinated. Consideration should be given to the diversity within ethnic and racial groups as relates to cultural practices and beliefs about vaccination. An appreciation that there may be varied reasons for refusing vaccination should lead to more particularized culturally sensitive interventions to successfully increase vaccination rates.

## Data Availability

The data presented in this study are available on request from the corresponding author.

## Supplementary Materials

The following are available online at www.mdpi.com/xxx/s1, Survey Instrument.

## Author Contributions

Conceptualization, N.M.G., J.E.L., S.Z., B.S., D.F., A.F.; methodology, N.M.G., J.E.L., S.Z., B.S., D.K., D.F., A.F.; software, N.M.G., D.M., J.E.L., D.K., S.Z., B.S.; validation, N.M.G., formal analysis, N.M.G., D.M., J.E.L., D.K.; resources, J.N., A.F.; data curation, S.Z., D.K.; writing—original draft preparation, N.M.G., A.F.; writing—review and editing, N.M.G., J.E.L., D.M., B.S., D.K., K.M., D.F., J.N., A.F.; visualization, N.M.G.; supervision, N.M.G., D.F., A.F. All authors have read and agreed to the published version of the manuscript.

## Informed Consent Statement

Informed consent was obtained from all subjects involved in the study.

## Data Availability Statement

The data presented in this study are available on request from the corresponding author.

## Acknowledgments

We thank employees of RUHS for their participation in this study.

