## Supplemental Materials for "Correlates of COVID-19 vaccine acceptance, hesitancy and refusal among employees of a safety net California county health system with an early and aggressive vaccination program: Results from a cross-sectional survey"

**Warmest greetings! Thank you for your service to RUHS patients. You are invited to complete a brief survey as a part of a research study. The aim of the study is to understand RUHS employees' views on COVID-19 vaccines. This study is being carried out by the Comparative Effectiveness and Clinical Outcomes Research Center (CECORC) at RUHS in collaboration with Claremont Graduate University (CGU).**

**Choice to do Survey**
**You have a choice to complete the survey or not. Even if you choose to participate, you can leave the survey at any time. Your answers to survey questions DO NOT affect your job status.**

**Confidentiality**
**Your participation stays anonymous. This means any answer that you provide will NOT be connected to you. This survey has been approved by the Research Subjects Protection Committee called the Institutional Review Board (IRB). Also, this survey DOES NOT collect any information that can be used to identify you.**

**Research Contact Information**
**If you have questions about the survey, please contact:**
**Anthony Firek, MD, Director of Medical Research**
**CECORC - RUHS 951-486-4489 and**

**Glen Moulton, MPH, MA(Ed) CIP, CCRP, Manager, Institutional Review Board**
**RUHS - 951-486-4098**

**General Survey Instructions**
**Choose your response(s) for each question. It is expected the survey will take less than 10 minutes to finish. PLEASE provide answers that are true to your own feelings and choices.**

**The survey has 3 parts:**

**PART 1. Demographics: In this part, questions about your age, gender, marital status, and other basic non-identifying information are included.**

**PART 2. Knowledge about COVID-19: This part includes questions about COVID-19 symptoms and disease.**

**PART 3. Views on COVID-19 Vaccines & Intent to Vaccinate: This part includes questions on your perspectives about COVID-19 vaccines.**

**Your participation in this survey will help in understanding the reasons why people accept or decline to be vaccinated.**

**By clicking “NEXT”, you agree (consent) to take this survey. We thank you for your participation in this important research.**

* 1. Please choose your age category.

- ◯ 18-29
- ◯ 30-49
- ◯ 50-64
- ◯ 65+

* 2. Please choose your gender.

- ◯ Female
- ◯ Male
- ◯ Non-binary/third gender
- ◯ Prefer not to answer
- ◯ Other

* 3. Please provide only the first 3 digits of your residence zip code.
*For example, if your zip code is* ***<u>923</u>****73 enter* ***923 only.***

* 4. What is your marital status?

- ◯ Single
- ◯ Married or civil union
- ◯ Divorced
- ◯ Separated
- ◯ Widowed/widower
- ◯ Living with partner

* 5. What is your race?

- ◯ American Indian or Alaska Native
- ◯ Asian
- ◯ Black or African American
- ◯ Native Hawaiian or Pacific Islander
- ◯ White
- ◯ Prefer not to answer
- ◯ Other (please specify)

* 6. What is your Ethnicity?

- ◯ Non-Hispanic/Non-Latino
- ◯ Hispanic/Latino
- ◯ Prefer not to say
- ◯ Other (please specify)

* 7. What is the highest degree you have received? (If you're currently enrolled in school, please indicate the highest degree you have received.)

- ◯ Less than a high school diploma
- ◯ High school degree or equivalent (e.g. GED)
- ◯ Some college, no degree
- ◯ Trade / Technical / Vocational Training
- ◯ Associate degree (e.g. AA, AS)
- ◯ Bachelor's degree (e.g. BA, BS)
- ◯ Master's degree (e.g. MA, MS, MEd)
- ◯ Professional degree (e.g. MD, DDS, DVM)
- ◯ Doctorate (e.g. PhD, EdD)
- ◯ Other (please specify)

* 8. Which of the following best describes your current work at RUHS?

- ◯ Nurse
- ◯ Doctor
- ◯ Laboratory, respiratory therapy, radiology personnel
- ◯ Administration or non-direct clinical support
- ◯ Maintenance, housekeeping, or dietary
- ◯ Pharmacist
- ◯ Medical Assistant
- ◯ Admissions and Collections Clerk
- ◯ Other (please specify)

* 9. In your current job, how often are you exposed to patients with COVID-19 on a weekly basis?

- ◯ I have no direct exposure (Example: working is an administrative office with no patient contact)
- ◯ I have minimal exposure (Example: working in a non-clinical position within the medical center OR within outpatient clinic)
- ◯ I have moderate exposure (Example: working in the ward on a non COVID ward or lab/ Radiology)
- ◯ I have a high exposure (Example: working in the COVID ward or ICU in any capacity)

* 10. What was the total combined income of your household in the past year?

- ◯ Less than $20,000
- ◯ $20,000 to $49,999
- ◯ $50,000 to $89,999
- ◯ $90,000 to $119,999
- ◯ $120,000 or above

* 11. How many people are currently living in your household, including yourself?

Total number of people:

Number of children (<18 years):

Number of adults (ages 18 - 64):

Number of seniors (over 65+):

* 12. Have you ever been told by a doctor or other health care provider that you have any of these health conditions?

| | Yes | No |
|---|---|---|
| Diabetes | ◯ | ◯ |
| Hypertension (High Blood Pressure) | ◯ | ◯ |
| Asthma | ◯ | ◯ |
| Serious heart conditions | ◯ | ◯ |
| Chronic lung disease | ◯ | ◯ |
| Chronic kidney disease | ◯ | ◯ |
| Liver disease | ◯ | ◯ |
| Weakened immune system (immunocompromised) | ◯ | ◯ |

* 13. What is your height?

Feet

Inches

* 14. What is your weight in pounds (lbs)?

**The vaccine against COVID-19 is now available. Personnel at RUHS are in the process of receiving vaccines on a tiered schedule.**

* 15. Please answer the following questions marking True, False, or I do not know.

| | True | False | I do not know |
|---|---|---|---|
| On average it takes 5–6 days from when someone is infected with COVID-19 for symptoms to show, however it can take up to 14 days | ◯ | ◯ | ◯ |
| People over the age of 65 are at greater risk of severe illness if they get COVID-19 | ◯ | ◯ | ◯ |
| COVID-19 can be spread from person to person. | ◯ | ◯ | ◯ |
| People with COVID-19 always show symptoms of being sick | ◯ | ◯ | ◯ |
| Most people who get COVID-19 only have mild symptoms | ◯ | ◯ | ◯ |
| I do not have to wear a mask after getting vaccinated. | ◯ | ◯ | ◯ |

* 16. Which of the following are common symptoms of COVID-19? Check all that apply.

- ☐ Fever
- ☐ Cough
- ☐ Shortness of breath
- ☐ Sore throat
- ☐ Runny or stuffy nose
- ☐ Muscle or body aches
- ☐ Headaches
- ☐ Fatigue (feeling tired)
- ☐ Diarrhea
- ☐ Loss of smell or taste
- ☐ Rash on face
- ☐ Blurry vision
- ☐ Earache
- ☐ Hair loss

* 17. Have you gotten a vaccine against COVID-19?

- ◯ Yes, one dose skip to question 20
- ◯ Yes, two doses skip to question 20
- ◯ No skip to question 18

* 18. When the opportunity comes to receive a vaccine for COVID-19, either through RUHS or from an outside facility, will you be vaccinated?

- Yes skip to question 21
- No skip to question 19
- Unsure skip to question 19

* 19. If you choose not to be vaccinated now, will you consider vaccination at a later date?

- ◯ Yes skip to question 21
- ◯ No skip to question 22
- ◯ Unsure skip to question 21

* 20. Thinking back to before you were vaccinated, please indicate whether the following influenced or would have influenced your decision to get vaccinated.

| | Definitely would not | Probably would not | Not sure | Probably would | Definitely would |
|---|---|---|---|---|---|
| If I had received a financial incentive | ○ | ○ | ○ | ○ | ○ |
| If I were entered in a raffle to win a gift card | ○ | ○ | ○ | ○ | ○ |
| If I had paid time off to get the vaccine | ○ | ○ | ○ | ○ | ○ |
| If getting vaccinated was a requirement for my job | ○ | ○ | ○ | ○ | ○ |
| If I believe I am healthy and can withstand a COVID infection | ○ | ○ | ○ | ○ | ○ |
| If I feel confident I can prevent COVID infection by using current precautions | ○ | ○ | ○ | ○ | ○ |
| If vaccination was promoted in my social media network | ○ | ○ | ○ | ○ | ○ |
| If I was convinced that getting vaccinated helped protect vulnerable members of my family or my community. | ○ | ○ | ○ | ○ | ○ |
| If someone I knew got sick, was hospitalized, died from COVID-19 | ○ | ○ | ○ | ○ | ○ |
| If colleagues or family members encouraged me to be vaccinated | ○ | ○ | ○ | ○ | ○ |
| If I knew that the pharmaceutical industry was not taking advantage of me | ○ | ○ | ○ | ○ | ○ |
| If my religious leaders said I should get vaccinated | ○ | ○ | ○ | ○ | ○ |
| If a trusted health care worker told me to get vaccinated | ○ | ○ | ○ | ○ | ○ |
| If I was sure that the vaccine is safe | ○ | ○ | ○ | ○ | ○ |
| If I was sure that the vaccine is effective and see people that were vaccinated not get sick with COVID-19 | ○ | ○ | ○ | ○ | ○ |
| If I believe there will be new medication to treat COVID infection soon | ○ | ○ | ○ | ○ | ○ |
| If getting vaccinated was required for me to attend social or sporting events or travel | ○ | ○ | ○ | ○ | ○ |

skip to question 23

* 21. Please indicate whether the following would have any influence on your decision to get vaccinated.

| | Definitely would not | Probably would not | Not sure | Probably would | Definitely would |
|---|---|---|---|---|---|
| If I were to receive a financial incentive | ○ | ○ | ○ | ○ | ○ |
| If I were entered in a raffle to win a gift card | ○ | ○ | ○ | ○ | ○ |
| If I were given paid time off to get the vaccine | ○ | ○ | ○ | ○ | ○ |
| If getting vaccinated was a requirement for my job | ○ | ○ | ○ | ○ | ○ |
| If I believe I am healthy and can withstand a COVID infection | ○ | ○ | ○ | ○ | ○ |
| If I feel confident I can prevent COVID infection by using current precautions | ○ | ○ | ○ | ○ | ○ |
| If vaccination was promoted in my social media network | ○ | ○ | ○ | ○ | ○ |
| If I was convinced that getting vaccinated helped protect vulnerable members of my family or my community. | ○ | ○ | ○ | ○ | ○ |
| If someone I knew got sick, was hospitalized, died from COVID-19 | ○ | ○ | ○ | ○ | ○ |
| If colleagues or family members encouraged me to be vaccinated | ○ | ○ | ○ | ○ | ○ |
| If I knew that the pharmaceutical industry was not taking advantage of me | ○ | ○ | ○ | ○ | ○ |
| If my religious leaders said I should get vaccinated | ○ | ○ | ○ | ○ | ○ |
| If a trusted health care worker told me to get vaccinated | ○ | ○ | ○ | ○ | ○ |
| If I was sure that the vaccine is safe | ○ | ○ | ○ | ○ | ○ |
| If I was sure that the vaccine is effective and see people that were vaccinated not get sick with COVID-19 | ○ | ○ | ○ | ○ | ○ |
| If I believe there will be new medication to treat COVID infection soon | ○ | ○ | ○ | ○ | ○ |
| If getting vaccinated was required for me to attend social or sporting events or travel | ○ | ○ | ○ | ○ | ○ |

skip to question 23

* 22. Imagine you are considering vaccination, please indicate whether the following would influence your decision to get vaccinated.

| | Definitely would not | Probably would not | Not sure | Probably would | Definitely would |
|---|---|---|---|---|---|
| If I were to receive a financial incentive | ◯ | ◯ | ◯ | ◯ | ◯ |
| If I were entered in a raffle to win a gift card | ◯ | ◯ | ◯ | ◯ | ◯ |
| If I were given paid time off to get the vaccine | ◯ | ◯ | ◯ | ◯ | ◯ |
| If getting vaccinated was a requirement for my job | ◯ | ◯ | ◯ | ◯ | ◯ |
| If I believe I am healthy and can withstand a COVID infection | ◯ | ◯ | ◯ | ◯ | ◯ |
| If I feel confident I can prevent COVID infection by using current precautions | ◯ | ◯ | ◯ | ◯ | ◯ |
| If vaccination was promoted in my social media network | ◯ | ◯ | ◯ | ◯ | ◯ |
| If I was convinced that getting vaccinated helped protect vulnerable members of my family or my community. | ◯ | ◯ | ◯ | ◯ | ◯ |
| If someone I knew got sick, was hospitalized, died from COVID-19 | ◯ | ◯ | ◯ | ◯ | ◯ |
| If colleagues or family members encouraged me to be vaccinated | ◯ | ◯ | ◯ | ◯ | ◯ |
| If I knew that the pharmaceutical industry was not taking advantage of me | ◯ | ◯ | ◯ | ◯ | ◯ |
| If my religious leaders said I should get vaccinated | ◯ | ◯ | ◯ | ◯ | ◯ |
| If a trusted health care worker told me to get vaccinated | ◯ | ◯ | ◯ | ◯ | ◯ |
| If I was sure that the vaccine is safe | ◯ | ◯ | ◯ | ◯ | ◯ |
| If I was sure that the vaccine is effective and see people that were vaccinated not get sick with COVID-19 | ◯ | ◯ | ◯ | ◯ | ◯ |
| If I believe there will be new medication to treat COVID infection soon | ◯ | ◯ | ◯ | ◯ | ◯ |
| If getting vaccinated was required for me to attend social or sporting events or travel | ◯ | ◯ | ◯ | ◯ | ◯ |

skip to question 23

* 23. What is your opinion about the following statements?

| | Strongly Disagree | Disagree | Neutral | Agree | Strongly Agree |
|---|---|---|---|---|---|
| Vaccines are important for my health. | ◯ | ◯ | ◯ | ◯ | ◯ |
| Getting the vaccine is important to protect the health of my family. | ◯ | ◯ | ◯ | ◯ | ◯ |
| Getting vaccines is a good way to protect myself from disease | ◯ | ◯ | ◯ | ◯ | ◯ |
| Overall, vaccines are safe/beneficial | ◯ | ◯ | ◯ | ◯ | ◯ |
| Overall, vaccines are effective | ◯ | ◯ | ◯ | ◯ | ◯ |
| Getting vaccinated is important for the health of others in my community | ◯ | ◯ | ◯ | ◯ | ◯ |
| The information I receive about vaccines from public health authorities/ my healthcare provider is reliable and trustworthy. | ◯ | ◯ | ◯ | ◯ | ◯ |
| Generally, I do what my doctor or health care provider recommends about vaccines for myself and my family. | ◯ | ◯ | ◯ | ◯ | ◯ |
| New vaccines carry more risk that older vaccines | ◯ | ◯ | ◯ | ◯ | ◯ |
| I am concerned about serious negative effects of vaccines | ◯ | ◯ | ◯ | ◯ | ◯ |
| I do not need vaccines for diseases that are not common anymore | ◯ | ◯ | ◯ | ◯ | ◯ |

* 24. Do you get the Flu vaccine every year as recommended?

- ◯ Yes skip to question 28
- ◯ No skip to question 27
- ◯ I skip some years skip to questions 25 & 26

* 25. If you do get the flu vaccine, please rank the following reasons for vaccination in order of importance. With #1 being the <u>most important</u> and #4 being the <u>least important.</u>

- [ ] I do not want to infect my family members
- [ ] I do not want to infect my coworkers or patients
- [ ] I do not want to become sick with the flu
- [ ] It is a requirement for employment

* 26. If you do NOT get the flu vaccine, please rank the following reasons for NOT vaccinating in order of importance. With #1 being the most important and #5 being the least important.

I do not believe vaccines are safe

☐ N/A

I am healthy and have a low risk for getting the flu

☐ N/A

I do not believe the flu vaccine is very effective

☐ N/A

I believe you can get the flu from the vaccine

☐ N/A

I have an allergic reaction to the vaccine

☐ N/A

skip to question 29

* 27. Please rank the reasons for NOT getting the flu vaccine, in order of importance. With #1 being the most important and #5 being least important.

I do not believe vaccines are safe

☐ N/A

I am healthy and have a low risk for getting the flu

☐ N/A

I do not believe the flu vaccine is very effective

☐ N/A

I believe you can get the flu from the vaccine

☐ N/A

I have an allergic reaction to the vaccine

☐ N/A

skip to question 29

* 28. Please rank the following reasons for flu vaccination in order of importance. With #1 being the most important and #4 being the least important.

I do not want to infect my family members

I do not want to infect my coworkers or patients

I do not want to become sick with the flu

It is a requirement for employment

skip to question 29

* 29. If you have children, do you feel it is important to have them vaccinated against childhood diseases?

- No
- Yes
- Not sure
- I don't have children

* 30. Have you or anyone you know tested <u>positive</u> for COVID-19?

- Yes skip to question 31
- No skip to question 32

* 31. Of those people who you know who tested positive for COVID-19, please indicate their level of symptoms

| | Symptoms |
|---|---|
| Myself | |
| Immediate family member | |
| Extended family member | |
| Friend who does not live with me | |
| Roommate or friend who lives with me | |
| Coworker | |
| Distant friend or a friend of a friend | |

* 32. How has the COVID-19 pandemic affected...

| | Severely decreased | Decreased | No effect | Improved | I do not know |
|---|---|---|---|---|---|
| My employment/income? | ◯ | ◯ | ◯ | ◯ | ◯ |
| My family's employment/income? | ◯ | ◯ | ◯ | ◯ | ◯ |
| My mental health and well being? | ◯ | ◯ | ◯ | ◯ | ◯ |
| My physical health? | ◯ | ◯ | ◯ | ◯ | ◯ |
| My ability to carry out my normal activities? | ◯ | ◯ | ◯ | ◯ | ◯ |

* 33. Please rate the following sources you use to get information about COVID-19.

| | Trustworthy | Neutral | Not Trustworthy |
|---|---|---|---|
| My primary doctor | ◯ | ◯ | ◯ |
| **CDC** (Center for Disease Control and Prevention), **WHO** (World Health Organization), local Department of Health | ◯ | ◯ | ◯ |
| News from the television or newspapers | ◯ | ◯ | ◯ |
| Friends | ◯ | ◯ | ◯ |
| Social Media (Facebook, Instagram, Twitter, etc) | ◯ | ◯ | ◯ |
| Celebrities/public figures | ◯ | ◯ | ◯ |
| Religious leaders | ◯ | ◯ | ◯ |
| Political leaders | ◯ | ◯ | ◯ |
| Family Members | ◯ | ◯ | ◯ |

* 34. Are you willing to answer a few more questions about your political beliefs?

- ◯ Yes skip to question 35
- ◯ No will End Survey

* 35. Please select the option that best describes your political ideology

- Very conservative
- Somewhat conservative
- Neither conservative nor liberal
- Somewhat liberal
- Very liberal

* 36. Who did you vote for in the last presidential election?

- Joe Biden
- Donald Trump
- Other / did not vote

END OF SURVEY
